# Age-Related Differences in the Choroid Plexus Structural Integrity Are Associated with Changes in Cognition

**DOI:** 10.1101/2025.02.27.25323022

**Authors:** Zhaoyuan Gong, Angelique de Rouen, Nathan Zhang, Joseph S.R. Alisch, Murat Bilgel, Yang An, Jonghyun Bae, Noam Y. Fox, Alex Guo, Susan M. Resnick, Caio Mazucanti, Samuel Klistorner, Alexander Klistorner, Josephine M. Egan, Mustapha Bouhrara

## Abstract

The choroid plexus (CP) plays a critical role in maintaining central nervous system (CNS) homeostasis, producing cerebrospinal fluid, and regulating the entry of specific substances into the CNS from blood. CP dysfunction has been implicated in various neurological and psychiatric disorders, including Alzheimer’s disease, Parkinson’s disease, and multiple sclerosis. This study investigates the relationship between CP structural integrity and cognitive decline in normative aging, using structural and advanced magnetic resonance imaging techniques, including CP volume, diffusion tensor imaging indices (mean diffusivity, MD, and fractional anisotropy, FA) and relaxometry metrics (longitudinal, T_1_, and transverse, T_2_, relaxation times). Our results show that diminished CP microstructural integrity, as reflected by higher T_1_, T_2_, and MD values, or lower FA values, is associated with lower cognitive performance in processing speed and fluency. Notably, CP microstructural measures demonstrated greater sensitivity to cognitive decline than macrostructural measures, i.e. CP volume. Longitudinal analysis revealed that individuals with reduced CP structural integrity exhibit steeper cognitive decline over time. Furthermore, structural equation modeling revealed that a latent variable representing CP integrity predicts faster overall cognitive decline, with an effect size comparable to that of age. These findings highlight the importance of CP integrity in maintaining cognitive health and suggest that a holistic approach to assessing CP integrity could serve as a sensitive biomarker for early detection of cognitive decline. Further research is needed to elucidate the mechanisms underlying the relationship between CP structural integrity and cognitive decline and to explore the potential therapeutic implications of targeting CP function to prevent or treat age-related cognitive deficits.

## Introduction

The choroid plexus (CP), a highly vascularized structure within the brain ventricles, is responsible for producing cerebrospinal fluid (CSF) by specialized epithelial cells. This process maintains the optimal composition and volume of CSF, providing essential cushioning and protection for the central nervous system (CNS) (1, 2). It also acts as a selective barrier between the blood and CSF, regulating the entry of substances into the CNS. Additionally, the CP structure facilitates bulk removal of waste products through the glymphatic system and plays pivotal roles in regulating chemical balance, ionic composition, immunosurveillance and influencing circadian rhythmicity (3–5). Dysregulation of these CNS processes can lead to cerebral tissue degeneration and neuroinflammatory responses, leading to disrupted neural signaling as well as concomitant cognitive impairments and motor dysfunction (6–8). Indeed, CP integrity is believed to play a significant role in cognition, as increasing research indicates that an enlarged CP is often associated with cognitive impairment in a myriad of conditions including mild cognitive impairment (9), Alzheimer’s disease (AD) (10, 11), Parkinson’s disease (PD) (12, 13), and multiple sclerosis (MS) (14, 15), where CP size correlates with the severity of cognitive decline. A larger CP may indicate a disruption in the brain’s clearance system, potentially contributing to cognitive deficits (16, 17). Therefore, measuring macrostructural and microstructural changes of CP through advanced neuroimaging techniques - notably magnetic resonance imaging (MRI) - could potentially develop imaging biomarkers for early detection of cognitive decline. However, there is a paucity of research investigating the relationship between CP structural health and cognitive impairment in normative aging, particularly from a longitudinal perspective. Importantly, microstructural changes in the CP are likely to precede macrostructural changes (*i.e*., volume changes) by decades, making it crucial to investigate microstructural alterations in relation to cognitive changes. Elucidating the role of the CP structural integrity in cognition could lead to the development of novel therapeutic strategies aimed at improving its function, offering promising avenues for early intervention and prevention of cognitive decline.

Established quantitative MRI (qMRI) techniques, including diffusion tensor imaging (DTI) metrics (mean diffusivity, MD, and fractional anisotropy, FA) and relaxometry indices (longitudinal relaxation time, T_1_, and transverse relaxation time, T_2_), enable assessment of differences in CP microstructural integrity with aging, obesity, and markers of neuroinflammation and AD pathology (18–22). DTI metrics offer a sensitive tool for monitoring CP changes in aging and elucidating the interplay between CP function, metabolic dysfunctions, and AD progression. DTI is sensitive to the underlying microarchitectural status of the CNS tissue and the degree and direction of water molecule mobility (23), and has been extensively used to study brain maturation and degeneration (24). Reduced FA is associated with structural degeneration, whereas an increase in MD reflects damage to cerebral tissue microstructural integrity with increased water mobility. Similarly, T_1_ and T_2_ both depend on macromolecular tissue composition, biochemistry and water mobility (25). Changes in T_1_ or T_2_ are directly associated with cerebral microstructural tissue changes, where higher values indicate advanced cerebral tissue deterioration (25). Notably, these associations between qMRI metrics and cerebral microstructural changes are also observed in CP (19), reinforcing the potential of qMRI for monitoring CP microstructural alterations. While studies using structural MRI have revealed that greater CP volume is inversely correlated with cognitive performance including in the AD continuum (9, 26), studies investigating microstructural differences in CP and cognition, as probed using DTI or relaxometry metrics, are lacking, including in normative aging. Yet, such investigations would further our understanding of the complex relationships between CP microstructure, function, and cognitive health (22).

This study aims to investigate the relationship between CP structural integrity, cognitive function, and aging, leveraging both structural and qMRI techniques. Specifically, we aim to examine CP microstructural differences associated with normative aging and longitudinal cognitive decline, clarify the link between CP microstructure and age-related cognitive health, and assess the utility of DTI and relaxometry metrics as biomarkers for early cognitive decline detection. This work advances our understanding of the complex interplay between CP structure and cognitive health.

## Materials and methods

### Participants

The MRI protocol received approval from the MedStar Research Institute and the National Institutes of Health (NIH) Intramural Ethics Committees. All examinations adhered to the standards set by the NIH Institutional Review Board. Study participants were recruited from the Baltimore Longitudinal Study of Aging (BLSA) and the Genetic and Epigenetic Signatures of Translational Aging Laboratory Testing (GESTALT) study (27, 28). Details regarding the study population, experimental design, and measurement protocols of BLSA and GESTALT have been described in previous publications (27, 28). The BLSA is a longitudinal cohort study initiated in 1958, conducted and funded by the National Institute on Aging (NIA) Intramural Research Program (IRP). The BLSA enrolls healthy, community-dwelling adults without major chronic conditions or functional impairments. Similarly, the GESTALT study, launched in 2015, is also conducted and funded by the NIA IRP, with virtually identical inclusion and exclusion criteria. Participants in both studies underwent evaluations at the NIA’s clinical research unit, with exclusions based on metallic implants, neurological or medical disorders, and cognitive impairment, as assessed through a comprehensive battery of cognitive tests (29).

### Magnetic Resonance Imaging (MRI)

For each participant, the imaging protocol for longitudinal and transverse relaxation times (T_1_, T_2_) and DTI metrics (FA and MD) consisted of:

#### *T*_1_ and *T*_2_ mapping (30–33)

3D spoiled gradient recalled echo (SPGR) images were acquired with flip angles (FAs) of [2 4 6 8 10 12 14 16 18 20]°, echo time (TE) of 1.37 ms, repetition time (TR) of 5 ms, and acquisition time of ∼5 min. Additionally, 3D balanced steady-state free precession (bSSFP) images were acquired with FAs of [2 4 7 11 16 24 32 40 50 60]°, TE of 2.8 ms, TR of 5.8 ms, and acquisition time of ∼6 min. To account for off-resonance effects, bSSFP images were acquired with radiofrequency excitation pulse phase increments of 0 or π (34). All SPGR and bSSFP images had an acquisition matrix of 150 × 130 × 94, voxel size of 1.6 mm × 1.6 mm × 1.6 mm. The double-angle method (DAM) was used to correct for excitation radiofrequency inhomogeneity (35). For this, two fast spin-echo images were acquired with FAs of 45° and 90°, TE of 102 ms, TR of 3000 ms, acquisition voxel size of 2.6 mm × 2.6 mm × 4 mm, and acquisition time of ∼4 min. All images were acquired with field of view (FoV) of 240 mm × 208 mm × 150 mm.

#### FA and MD mapping (24, 36)

DTI protocol consisted of diffusion-weighted images (DWI) acquired with single-shot Echo Planar Imaging (EPI), TR of 10,000 ms, TE of 70 ms, two b-values of 0 and 700 s/mm^2^, with the latter encoded in 32 directions. The acquisition matrix was 120 × 104 × 75, with voxel size of 2 mm × 2 mm × 2 mm, with FoV of 240 mm × 208 mm × 150 mm.

MRI scans were performed on a 3T whole-body Philips MRI system (Achieva, Philips Healthcare, Best, The Netherlands), utilizing the internal quadrature body coil for transmission and an eight-channel phased-array head coil for signal reception.

### MRI processing

MRI processing and analysis details can be found in Reference (19). In brief, scalp and nonparenchymal regions were eliminated using FSL. CP volume was calculated using FreeSurfer for each participant. CP masks were thoroughly examined and corrected manually when needed. T_1_ and T_2_ mapping as well as FA and MD mapping, were performed using FSL and in-house MATLAB scripts. Mean T_1_, T_2_, FA, and MD values were extracted from the CP region of interest (ROI) using the CP mask from FreeSurfer.

### Cognitive assessment

Cognitive domain scores were obtained for memory (California Verbal Learning Test (37) immediate and long-delay free recall), attention (Trail Making Test (38) Part A and Digit Span (39) Forward), executive function (Trail Making Test Part B and Digit Span Backward), verbal fluency (Category (40) and Letter Fluency (41)), and processing speed (Trail Making Test (38) Part A and Digit Symbol Substitution Test). To obtain domain scores, each test score was first converted to a z-score using the baseline mean and standard deviation, and these z-scores were averaged within each cognitive domain. Before computing the z-scores for Trail Making Test Parts A and B, the individual cognitive test scores (time to completion, in seconds) were log-transformed and negated so that higher z-scores indicated shorter time to completion.

### Statistical analyses

We conducted three statistical analyses to assess the relationship between CP and cognition. All reported p-values in the main manuscript are adjusted for multiple comparisons across all regressions using the Benjamini & Hochberg procedure. Analyses were programmed in R 4.4.2, with lme4, lavaan, and tidyverse packages.

#### CP Structural Integrity vs. Cognition

To assess the cross-sectional relationship between CP structural integrity and cognition, we paired cognition scores with the closest MRI scan in time to create cross-sectional datasets. We then performed multiple linear regressions to evaluate the association between each MRI metric and cognitive domain, specified as:

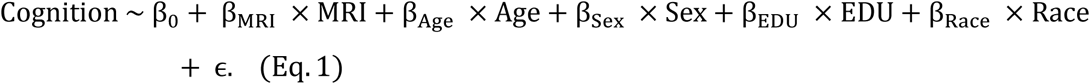

where Cognition represents one of five cognitive domains: processing speed (PS), memory (MEM), verbal fluency (FLU), executive function (EF), or attention (ATT). MRI metrics include CP volume (Vol), longitudinal relaxation time (T_1_), transverse relaxation time (T_2_), fractional anisotropy (FA), and mean diffusivity (MD). We observed collinearity between MRI metrics and Age, which weakened the significance of expected age-related cognitive declines. To address this, we regressed MRI metrics on covariates and replaced the original MRI values with their residuals. Covariates included Age (at the time of MRI), Sex (coded with male as the reference group), EDU (years of education), and Race (categorized as White, Black, or Other, with White as the reference group). All continuous variables were standardized for analysis.

#### CP Structural Integrity vs. Changes in Cognition

To investigate the relationship between CP integrity and longitudinal cognitive changes, we constructed longitudinal datasets by pairing repeated cognitive assessments with each participant’s single MRI scan. The longitudinal dataset comprised 320 observations from 116 unique participants. We employed linear mixed-effects models:

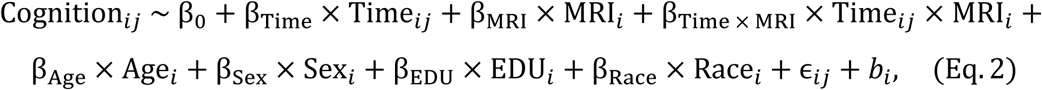

where Cognition*_ij_* represents the cognitive score for subject *i* at time point *j* in PS, MEM, FLU, EF, or ATT. Time*_ij_* is the Time-to-MRI calculated as the time difference between the time point *j* and the time at MRI scan for subject *i*. MRI*_i_* denotes the MRI metric for subject *i* at the time of the

MRI scan. The interaction term Time*_ij_*×MRI*_i_* tests whether MRI metrics modulate the rate of cognitive change over time. *b_i_* is the random intercept for subject *i*, and ɛ*_ij_* is the residual error for subject *i* at time point *j*. Age*_i_*, Sex*_i_*, EDU*_i_*, and Race*_i_* were included as covariates, as previously defined.

#### Overall CP Integrity and Overall Cognitive Function

We employed structural equation modeling (SEM) to investigate the relationships between latent constructs representing CP integrity and cognitive decline. SEM is a comprehensive statistical approach that facilitates the analysis of complex relationships involving both measured variables and unobserved constructs (latent variables). In this study, we opted to construct two comprehensive latent variables: CP Integrity (CPI) and Cognitive Function (CogFun). The CPI latent variable represents the unobservable overall structural integrity of the CP, inferred collectively from macrostructural and microstructural MRI measurements. In contrast, CogFun represents the unobservable overall rate of cognitive function changes, derived from all rates of change in the evaluated cognitive domains. By modeling these latent variables, we aimed to holistically assess the impact of CPI on overall cognitive function. The measurement model for CPI (exogenous latent variable) is defined as:

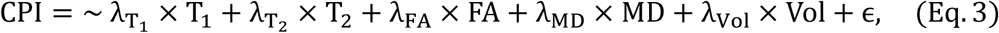

where λ represents factor loadings, and ɛ represents residual variance. Similarly, the measurement model for CogFun (endogenous latent variable) is defined as:

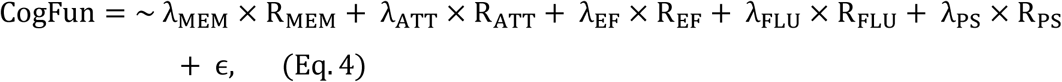

where R_MEM_, R_ATT_, R_EF_, R_FLU_, R_PS_ are rates of change in memory, attention, executive function, fluency, and processing speed, respectively. We modeled the structural relationship between CP Integrity and cognitive function as:

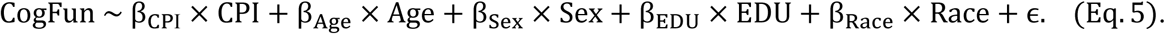

All results are presented in standardized form, with variables scaled to have a mean of zero and a standard deviation of one. This scaling allows parameter estimates (e.g., factor loadings, regression coefficients, and variances) to be interpreted as standardized effects.

## Results

Table 1 provides an overview of the study demographics. A total of 116 unique subjects were included. In the cross-sectional cohort, the median time between cognitive testing and MRI scanning was 0 years (mean: −0.17 years; SD: 0.6 years). In the longitudinal cohort, participants underwent an average of 2.76 cognitive assessments (SD: 2.74), with a median follow-up duration of 5.3 years (mean: 8.8 years; SD: 7.9 years). The average age at the time of MRI scanning was 57.2 years, and 44.8% of the participants were female. The cohort’s mean education level was 16.2 years (SD: 2.8 years). The cohort comprised 70.7% of white participants, indicating an imbalance in racial representation.

**TABLE 1.**
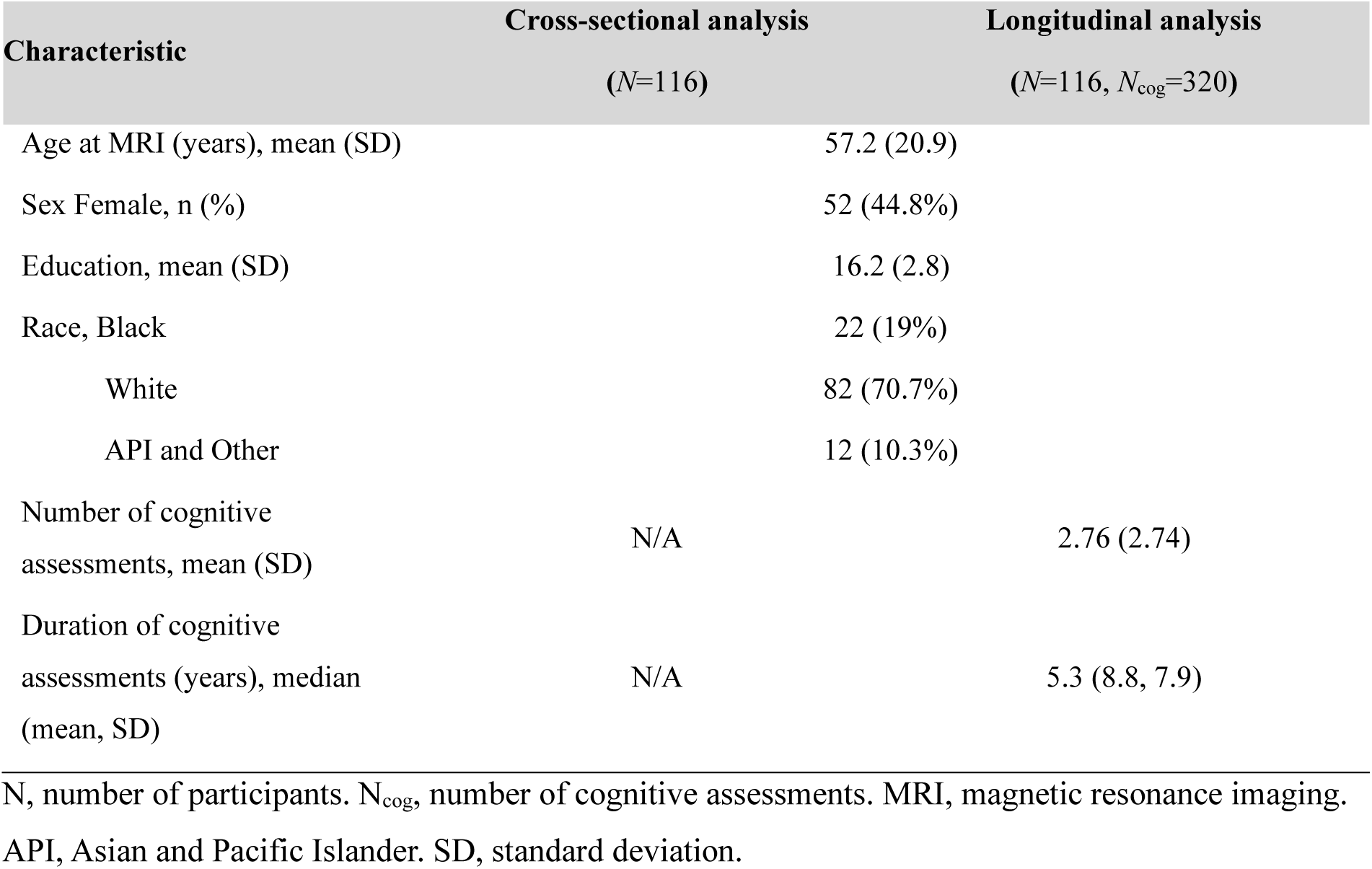
Participant demographics for cross-sectional and longitudinal analysis.

Figure 1 shows the results of the multiple linear regression analysis (Eq. 1) examining the cross-sectional relationship between each MRI metric (*i.e.*, CP volume, T_1_, T_2_, MD, and FA) and each cognitive domain (*i.e.*, PS, MEM, FLU, EF, and ATT). As expected, age was negatively associated with cognition in all examined regressions. These associations were all statistically significant. Examples of these associations are shown in Fig. 1B and C. Importantly, CP volume, T_1_, T_2_, and MD exhibited negative associations with cognition, while FA exhibited positive association with cognition, indicating that worse macrostructural CP integrity, represented by higher CP volume, and worse microstructural CP integrity, as measured by higher T_1_, T_2_, MD values and lower FA, are associated with worse cognition (Fig. 1D). However, these associations were statistically significant only for MD vs. FLU and PS as well as T_1_, T_2_, and FA vs. PS (Fig. 1D). Representative plots of these associations are shown in Fig. 1E and F. In addition, sex differences were significant, with males scoring higher in EF and ATT but lower in MEM. Higher education was significantly associated with better performance in MEM, FLU, EF, and ATT. White participants showed significantly better performance in ATT, EF, and PS. We note that the main purpose of this study is not to investigate the covariates, and the results may not be generalizable due to confounding factors. Complete analysis results are provided in the supplementary materials.

**Figure 1.**
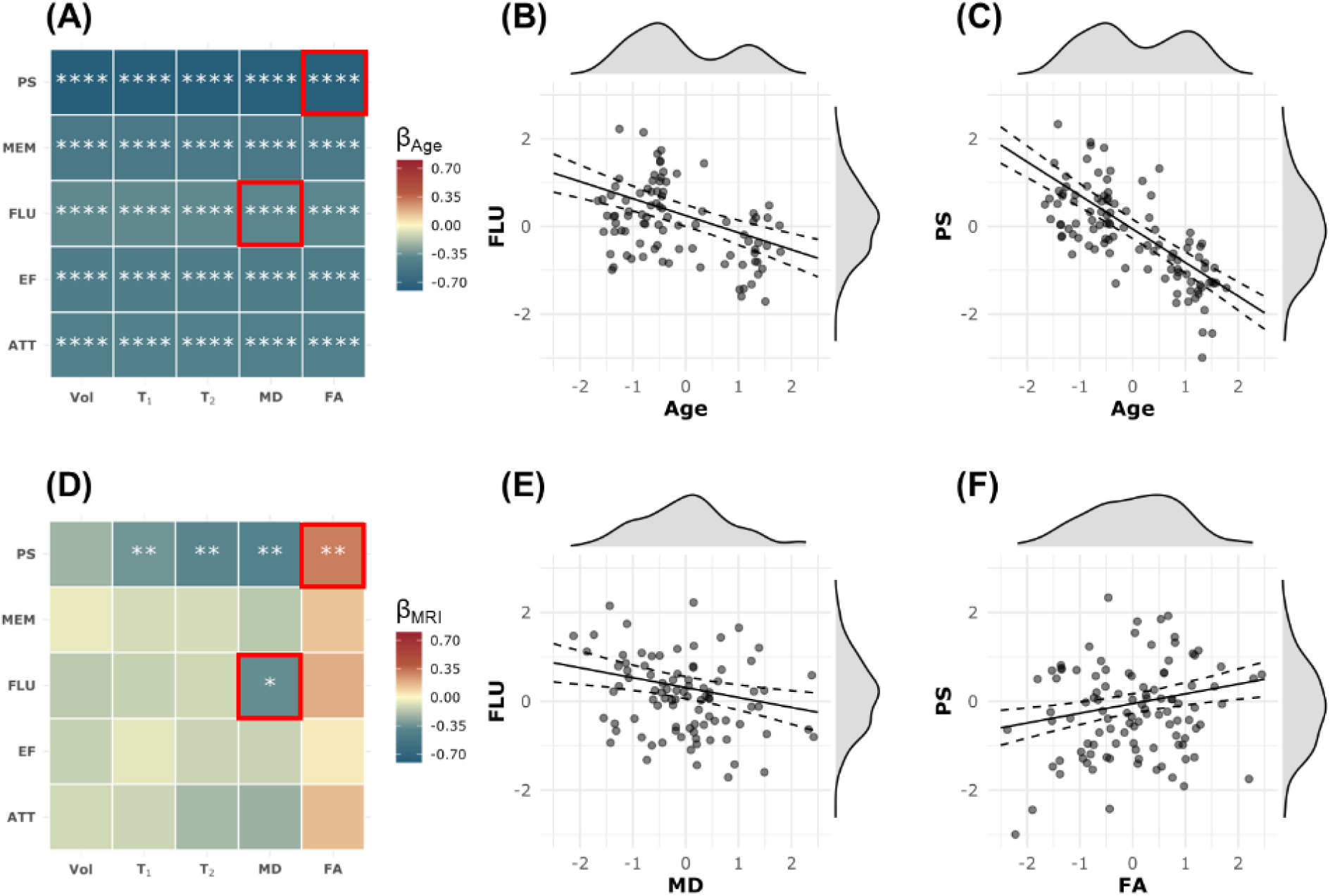
Pairwise multiple linear regression was conducted for each MRI metric and each cognitive domain. The explicit regression equation is: *Cognition* ∼ β_0_ + β_*MRI*_ × *MRI* + β_*Age*_ × *Age* + β_*Sex*_ × *Sex* + β_*EDU*_ × *EDU* + β_*Race*_ × *Race* + ∈, where Cognition includes processing speed (PS), memory (MEM), verbal fluency (FLU), executive function (EF), and attention (ATT). MRI metrics include the residuals of CP volume (Vol), T_1_ relaxation time (T_1_), T_2_ relaxation time (T_2_), fractional anisotropy (FA), and mean diffusivity (MD) after regressing with other covariates. All continuous variables are standardized. **Panels A and D** show the results for the regression coefficients of Age and MRI metrics, respectively. The estimated β coefficients are color-coded and statistically significant results are marked with asterisks, where * indicates p < 0.05, ** indicates p < 0.01, *** indicates p < 0.001, and **** indicates p < 0.0001. All p-values are corrected for multiple comparisons using the Benjamini–Hochberg (BH) procedure. **Panels B and E** present example results for the regression where the MRI metric is MD and the cognitive domain is FLU. There is a significant negative association between FLU and age (Panel B) and a significant negative association between MD and FLU (Panel E). High MD values indicate reduced microstructural integrity, which is associated with lower FLU. **Panels C and F** show example results for the regression where the MRI metric is FA and the cognitive domain is PS. In addition to a significant negative association between PS and age (Panel C), there is a significant positive association between FA and PS (Panel F). High FA values indicate greater microstructural integrity, which is associated with higher PS scores.

Figure 2 shows the results of the linear mixed-effects analysis (Eq. 2) examining the relationship between each MRI metric (*i.e.*, CP volume, T_1_, T_2_, MD, and FA) and longitudinal changes in each cognitive domain (*i.e.*, PS, MEM, FLU, EF, and ATT). Results are shown for the main parameters of interest relevant to this investigation, namely, MRI fixed effect, Time-to-MRI, and Time × MRI interaction. MRI metrics of CP volume, T_1_, T_2_, and MD exhibited negative associations with cognition, while FA exhibited a positive association with cognition, indicating that worse CP structural integrity is associated with lower cognitive performance. These associations reached statistical significance for MD vs. PS, FLU and ATT as well as T_1_, T_2_ and FA vs. PS (Fig. 2A). Examples of plots of these associations are shown in Fig. 2B and C. Further, as expected, Time-to-MRI exhibited overall negative association with cognition, reaching significance in various pairwise MRI metrics vs. cognitive domains as indicated in Fig. 2D-F. Importantly, the association between Time × MRI interaction and cognition exhibited negative associations for Vol, T_1_, T_2_ and MD and positive associations for FA (Fig. 2G). These associations were statistically significant for several MRI metrics vs. cognitive domains (Fig. 2G). These analyses indicate that lower CP structural integrity, as reflected by higher CP volume, T_1_, T_2_ and MD values or lower FA values, is associated with steeper longitudinal decline in cognition. Fig.2 H and I show the example of higher T_1_ value, corresponding to the value at the 75^th^ percentile marked with the green lines, is associated with steeper decline in PS and EF. Sex differences were significant, with males scoring higher in ATT but lower in MEM. Higher education was significantly associated with better performance in all cognitive domains, while White participants showed significantly better performance in ATT, EF, and PS. Same caution should be exercised for generalizing the results for covariates. Complete analysis results are provided in the supplementary materials.

**Figure 2.**
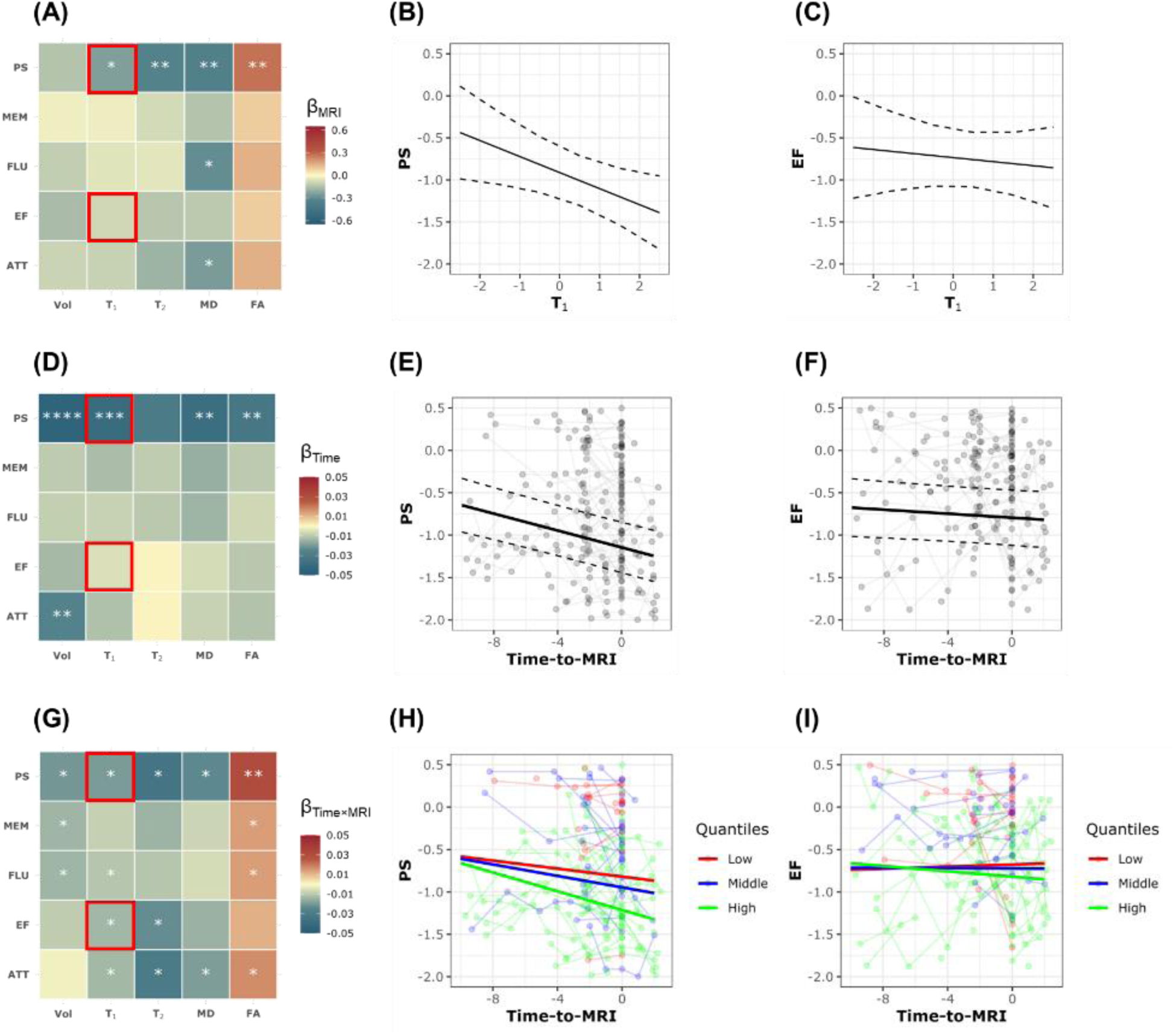
Pairwise linear mixed-effects (LME) models were conducted for each MRI metric and each cognitive domain. The explicit regression equation is: *Cognition*_*ij*_ ∼ β_0_ + β_*Time*_ × *Time*_*ij*_ + β_*MRI*_ × *MRI*_*i*_ + β_*Time* × *MRI*_ × *Time*_*ij*_ × *MRI*_*i*_ + β_*Age*_ × *Age*_*i*_ + β_*Sex*_ × *Sex*_*i*_ + β_*EDU*_ × *EDU*_*i*_ + β_*Race*_ × *Race*_*i*_ + ∈_*ij*_ + *b*_*i*_, where Cognition_ij_ includes longitudinal measurements of processing speed (PS), memory (MEM), verbal fluency (FLU), executive function (EF), and attention (ATT) for subject i at timepoint j. Time_ij_ represents the time to the only MRI scan of subject i at timepoint j. MRI metrics (MRI_i_) include CP volume (Vol), T_1_ relaxation time (T_1_), T_2_ relaxation time (T_2_), fractional anisotropy (FA), and mean diffusivity (MD) for subject i at the time of MRI scan. All continuous variables are standardized except for Time_ij_. **Panels A, D and G** show the results for the regression coefficients of MRI metrics, Time and Time × MRI interaction, respectively. The estimated β coefficients are color-coded and statistically significant results are marked with asterisks, where * indicates p < 0.05, ** indicates p < 0.01, *** indicates p < 0.001, and **** indicates p < 0.0001. All p-values are corrected for multiple comparisons using the Benjamini–Hochberg (BH) procedure. **Panels B, E, and H** present example results for the LME model where the MRI metric is T_1_ and the cognitive domain is PS. A significant negative association between the fixed effect of T_1_ and PS is observed (Panel B). This indicates that individuals with higher T_1_ values tend to have consistently lower cognitive scores over time. PS also shows a significant linear decline over time (Panel E). Most importantly, the significant negative interaction term indicates that individuals with higher T_1_ values experience faster declines in PS over time compared to those with lower T_1_ values (green line vs. red line in Panel H). **Panels C, F, and I** present example results for the LME model where the MRI metric is T_1_ and the cognitive domain is EF. No significant association is found between the fixed effect of T_1_ and EF (Panel C). EF also does not show a significant general decline over time (Panel F). However, a significant negative interaction term is observed, indicating that individuals with higher T_1_ values experience faster declines in EF over time compared to those with lower T_1_ values (green line vs. red line in Panel I). Increased T_1_ relaxation times could reflect microstructural changes such as reduced myelin integrity or axonal damage, which are known to impair cognitive processes and may exacerbate cognitive decline over time observed here.

Finally, Figure 3 shows the results of the SEM analysis investigating the relationship between combined MRI metrics (CPI) and combined change rates of cognitive function (CogFun) (Eqs. 3-5). As expected, derived rates of cognitive change exhibit decreased trends as a function of age, especially at older ages (Fig. 3A). Our analysis reveals that the measurement model for CP damage indicates significant standardized loadings for all MRI metrics. Interestingly, CP volume, a measure of macrostructural integrity, contributes the least to the latent variable as compared to all other MRI metrics, which are measures of microstructural alterations. Of note, FA exhibits a negative loading, as lower FA values reflect worse microstructural integrity. In the measurement model for cognitive changes, all rates of cognitive domain changes display significant loadings, with fluency contributing the least. The structural model reveals that CPI is negatively associated with CogFun (β = −0.41, p < 0.001), indicating that worse overall CP structural integrity predicts faster cognitive decline. This effect is close to that of age (β = −0.55, p < 0.001), highlighting a strong association. Furthermore, education exhibits a protective effect (β = 0.28, p = 0.001), whereas the effects of sex (β = −0.17, p = 0.045) and race (β = −0.06, p = 0.456) are weaker, with the latter being nonsignificant.

**Figure 3.**
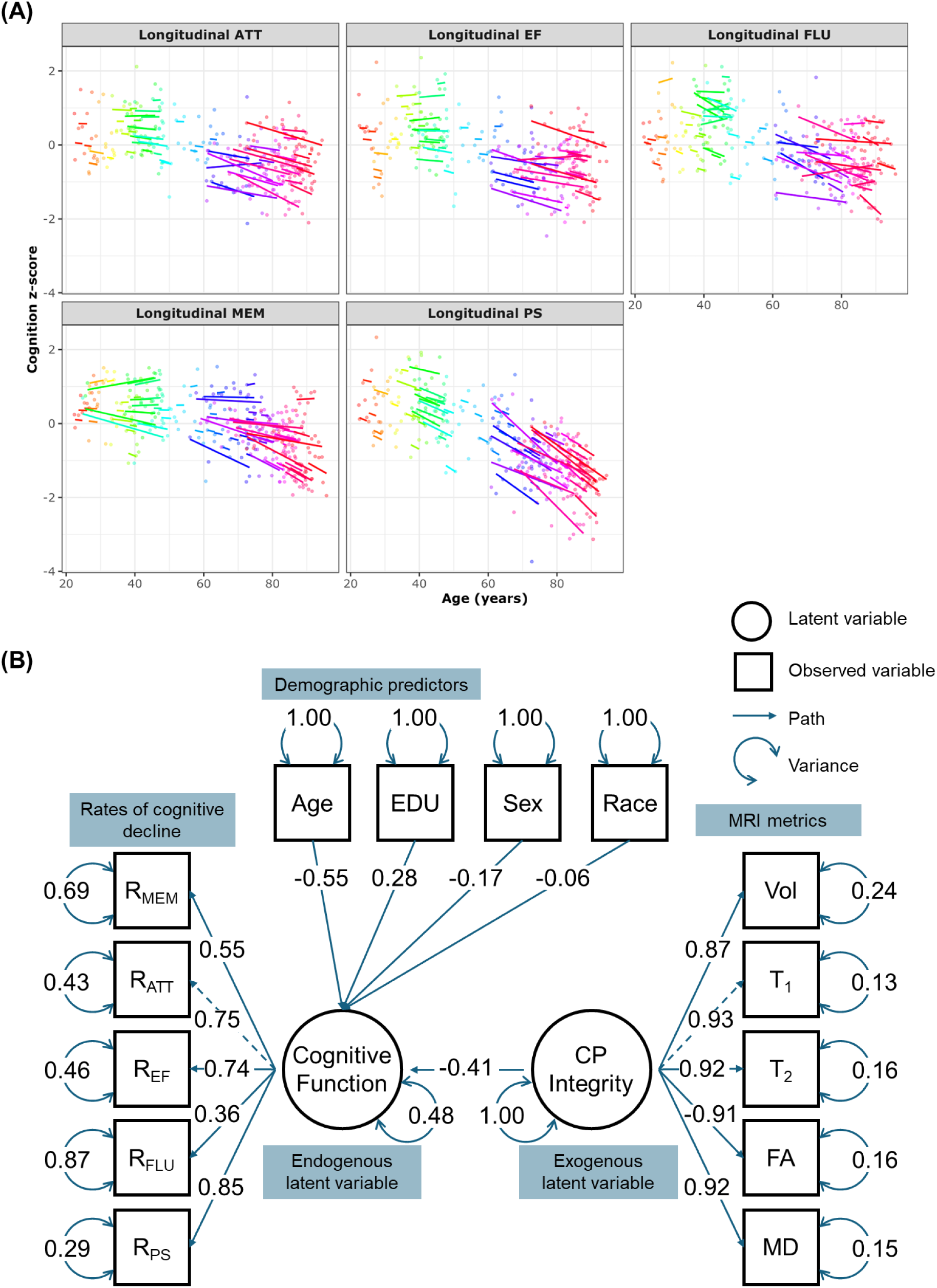
Structural equation modeling (SEM) was applied to investigate the relationship between combined MRI metrics (CPI) and combined rates of cognitive decline (CogFun). **Panel A** displays the estimated rates of Cognitive-decline for each subject, derived from linear mixed-effects models: *Cognition*_*ij*_ ∼ β_0_ + β_*Time*_ × *Time*_*ij*_+ *b*_*Time*,*i*_ × *Time*_*ij*_ + ∈_*ij*_ + *b*_*i*_. The rate of cognitive decline is defined as the sum of the fixed effect (β_Time_) and the subject-specific random effect (b_Time,i_). Each subject is color-coded and has a fitted decline line if more than two longitudinal measurements exist. **Panel B** visualizes the SEM results. The measurement model for CP integrity, CPI, (exogenous latent variable) is defined as: *CPI* = ∼ λ*T*_1_ × *T*_1_ + λ_*T*2_ × *T*_2_ + λ_*FA*_ × *FA* + λ_*MD*_ × *MD* + λ_*Vol*_ × *Vol* + ∈, where λ represents factor loadings, and ɛ represents residual variance. Similarly, the measurement model for Cognitive-function, Cog-Fun, (endogenous latent variable) is defined as: *CogFun* = ∼ λ_*MEM*_ × *R*_*MEM*_ + λ_*ATT*_ × *R*_*ATT*_ + λ_*EF*_ × *R*_*EF*_ + λ_*FLU*_ × *R*_*FLU*_ + λ_*PS*_ × *R*_*PS*_ + ∈, where R_MEM_, R_ATT_, R_EF_, R_FLU_, R_PS_ are rates of decline in memory, attention, executive function, fluency, and processing speed, respectively. The structural regression model is: *CogFun* ∼ β_*CPI*_ × *CPI* + β_*Age*_ × *Age* + β_*Sex*_ × *Sex* + β_*EDU*_ × *EDU* + β_*Race*_ × *Race* + ∈. The results are presented in the standardized form, allowing the parameter estimates to be interpreted as standardized effects. The measurement model for CPI indicates that all MRI metrics have significant standardized loadings, with CP volume contributing less to the latent variable because it reflects macrostructure, while other MRI metrics probe microstructure. Additionally, FA shows a negative loading because, unlike other MRI metrics, lower FA values indicate greater microstructural damage. In the measurement model for CogFun, all rates of cognitive domain changes have significant loadings, with fluency contributing the least. The structural model reveals that CPI has a significant negative effect on CogFun (β = −0.41, p < 0.001), suggesting that higher CPI predicts faster cognitive decline. This effect is comparable to that of age (β = −0.55, p < 0.001), indicating a strong association. Education (β = 0.28, p = 0.001) has a protective effect, while the effects of sex (β = −0.17, p = 0.045) and race (β = −0.06, p = 0.456) are weaker, with race being nonsignificant.

## Discussion

The present study revealed significant associations between CP integrity, as measured by MRI metrics, and cognitive function across various domains. Notably, our results showed that CP volume, T_1_, T_2_, and MD values were negatively correlated with cognitive performance, whereas FA values exhibited positive associations, indicating that compromised CP structural integrity is linked to poorer cognitive function. These findings were statistically significant for PS and FLU, suggesting that CP integrity plays a critical role in maintaining optimal cognitive performance. Furthermore, our analysis revealed that the associations between CP integrity and cognitive function were influenced by demographic factors, such as age, sex, education, and race, highlighting the importance of considering individual differences and sociological disparities related to demographic factors when examining the neural correlates of cognitive function. The observed relationships between CP integrity and cognitive decline also underscore the potential utility of MRI metrics as biomarkers for monitoring cognitive changes and detecting early signs of neurodegenerative diseases. Overall, our findings contribute to a deeper understanding of the complex interplay between brain structure and cognitive function and have significant implications for the development of novel diagnostic and therapeutic strategies aimed at promoting cognitive health and mitigating cognitive decline.

Existing evidence shows the sensitivity of microstructural changes to cognitive decline, which precedes macrostructural changes by decades (42, 43). By leveraging advanced MRI techniques, including diffusion and relaxometry, we demonstrate that microstructural alterations in the CP are more sensitive indicators of CP dysfunction and cognitive decline than macrostructural measurement. Previous research highlights the association between CP volume and cognitive impairment in various conditions. However, our study addresses a significant gap in the literature by investigating the relationship between CP microstructural integrity and cognitive decline in normative aging, using qMRI techniques. While previous studies have relied on structural MRI to examine the relationship between CP volume and cognitive performance (9, 10, 14, 44–47), our study provides novel insights into the microstructural changes underlying CP dysfunction and cognitive decline.

The mechanisms underlying the association between CP degeneration and cognitive decline are complex and multifaceted. Previous studies suggest that CP dysfunction may contribute to cognitive decline through various pathways, including disrupted CSF dynamics (48–50), neuroinflammation (51–53), and impaired glymphatic function (54–56). The age-related increase in CP volume may lead to an inflammatory response at the blood/CP-CSF barrier as well as in the brain itself. Furthermore, the CP’s role in regulating the entry of substances into the CNS and facilitating the removal of waste products from the brain may be compromised with aging (48, 49, 57, 58), leading to the accumulation of toxic substances and exacerbating cognitive decline. Additionally, the association between increased CP volume and impaired glymphatic function may also contribute to cognitive decline (54–56), as the glymphatic system plays a critical role in clearing metabolites from the brain. Further studies are needed to elucidate the underlying mechanisms and to determine the causal relationships between CP degeneration, neuroinflammation, and cognitive decline.

Although our study provides novel clinical insights into the relationship between CP structural integrity and cognitive decline in normative aging, some limitations should be acknowledged. First, our study employed a single time-point MRI assessment, which precludes the examination of longitudinal changes in CP structural integrity. Future studies incorporating longitudinal MRI assessments are necessary to verify the temporal relationships between CP structural changes and cognitive decline. Second, our study relied on indirect MRI markers of CP microstructural integrity, which cannot fully capture the complex cellular and molecular changes occurring within the CP. The development of more sensitive and specific biomarkers is needed to further elucidate the mechanisms underlying CP dysfunction and cognitive decline. Finally, our study did not control for potential confounding variables, such as sleep quality, physical activity, and social engagement, which may influence the relationship between CP microstructural integrity and cognitive decline. Future studies should strive to incorporate these variables to provide a more comprehensive understanding of the complex interplay between CP function, lifestyle factors, and cognitive health.

## Conclusions

Our findings demonstrate that compromised CP structural integrity is significantly associated with cognitive decline in aging among cognitively unimpaired individuals. This underscores the importance of CP health in maintaining cognitive function and suggests that advanced CP imaging biomarkers could aid in early detection and intervention strategies for cognitive decline.

## Data Availability

Availability of data and materials: The datasets generated and/or analyzed during this study are available upon reasonable request to the corresponding author, contingent upon institutional approval.

